# Economic crisis and post-COVID adaptations in Directly Observed Therapy practices among tuberculosis patients in Colombo, Sri Lanka: A cross-sectional study

**DOI:** 10.1101/2025.09.15.25335789

**Authors:** Tara Alahakoon, Ravindu Jayaweera, Dasha Asienga, Amy S. Wagaman, Alexandra E. Purdy, Onali B.W. Rajapakshe

## Abstract

**Background:** The World Health Organization strongly recommends the Directly Observed Therapy (DOT) strategy in managing tuberculosis (TB). In Sri Lanka, TB is anticipated to rise following the country’s economic crisis, particularly in urban Colombo. Family members have been appointed as Directly Observed Treatment providers since the COVID-19 pandemic. However, no study has yet evaluated the current status of DOT practices in Sri Lanka as affected by the country’s economic crisis and transition to family-administered DOT services. The objective of this study is to describe adherence to DOT practices and factors associated with these practices among TB patients in Colombo under this current system. It is the first to determine attitudes and practices related to DOT in Sri Lanka, as well as to highlight barriers and facilitators to treatment among its TB patients.

**Methods:** This descriptive cross-sectional study was conducted among 450 continuation phase patients of age ≥ 18 years registered at the Central Chest Clinic in Colombo. Data was collected via an interviewer-administered questionnaire and chart review. Multivariate and regression analyses were performed, with DOT compliance defined as having a DOT provider.

**Results:** Of 450 patients, 117 (26.0%) patients were DOT compliant, and most DOT noncompliant patients resorted to self-administered therapy (SAT). Nevertheless, DOT or lack thereof had no statistically significant effect on clinical outcomes including sputum conversion, change in BMI, and missed doses. Factors associated (α < 0.05) with noncompliance were male sex, loss of income due to diagnosis, lack of physical disability, inadequate social support, and awareness of curability. Patients aware that a DOT provider is meant to help them take their medication were more likely to be compliant.

**Conclusion:** Provided sufficient health education and socioeconomic support, SAT may be a feasible DOT alternative in resource-limited circumstances in which facility-based DOT is not attainable.

## Introduction

Despite the availability of curative treatment, tuberculosis (TB) remains a widely prevalent infectious disease and global health challenge. Strongly associated with poverty, TB is further exacerbated by malnutrition, overcrowding, poor ventilation, and stigma of diagnosis.^1^ Moreover, the COVID-19 pandemic has had lasting effects on TB management, disrupting health services, limiting treatment access, and stalling or reversing prior progress towards the World Health Organization (WHO)’s End TB targets.^2^

TB remains the second most prevalent infectious disease in Sri Lanka, with 9180 cases and 780 deaths (death rate = 8.2%) reported in 2024 and 2023, respectively (personal communication, National Programme for Tuberculosis Control and Chest Diseases (NPTCCD)). Colombo, the most urbanized district, consistently reports the highest TB burden, with 2218 cases in 2024 and 160 deaths in 2023 (personal communication, NPTCCD). Malnutrition is by far the nation’s largest risk factor, followed by alcoholism, smoking, diabetes, and HIV.^3^ Additionally, Sri Lanka’s ongoing economic crisis has increased poverty and malnutrition, particularly among Colombo’s urban poor, suggesting an imminent increase in communicable diseases such as TB.^4–8^

Endorsed by the WHO in 1993, the Directly Observed Therapy (DOT) strategy requires the patient’s ingestion of anti-TB drugs under the supervision of a trusted witness–a method credited with treatment success worldwide.^9^ By improving adherence to the lengthy treatment regimen, DOT minimises transmission.^9^ In contrast, poor DOT compliance is a key contributor to treatment failure, delayed sputum conversion, relapse rates, and increased drug resistance, thereby increasing the severity of the public health threat.^9^

The NPTCCD is the Ministry of Health’s central organisation responsible for TB control throughout Sri Lanka. Its network consists of 26 District Chest Clinics (DCC), under which field-level primary healthcare facilities have provided DOT services since 1997.^10^ DOT was fully implemented within the country from 2010 to pre-COVID times.^10^ However, due to the COVID-19 pandemic, DOT services diminished to nearly nonexistent levels in districts including Colombo and responsibility was entrusted to family members.^11^ Although the pandemic has subsided, DOT centres have not been re-established.^11^ With more responsibility for their health now delegated to the patient, alternatives to DOT are increasingly necessary.

This study aims to identify the prevalent knowledge, attitudes, and socioeconomic factors affecting the ability of TB patients in Colombo, Sri Lanka to engage in DOT practices. It seeks to collect information on the extent of adherence to anti-TB treatment post-COVID-19, of which there is none so far. In addition, clinical outcomes of patients’ alternate treatment practices were assessed to compare their efficacy. By elucidating the gaps in service delivery, informed policy changes can address the roots of poor DOT compliance and develop solutions that are both effective and socioeconomically feasible.

## Methods

### Study setting

The study was conducted in the district of Colombo, a cosmopolitan coastal city and the economic and cultural hub of Sri Lanka. Although the country suffers shortages of essential drugs and healthcare centres restrict free distribution of medicines, government commitment and international financial support keep TB treatment drugs readily available.^5,12^

A Central Chest Clinic (CCC) and three branch clinics open six days a week serve the Colombo district’s entire population of 2.3 million.^13^ This study was performed at the CCC, which is located within the district’s most population-dense area – a known TB hotspot. At the CCC, case detection, initiation of anti-TB treatment, and follow-up services are provided free of charge by the Ministry of Health. Following each visit, a patient receives either a week’s worth of anti-TB drugs during the intensive phase or a month’s worth during the continuation phase.^11^

### Study sample and data collection

This descriptive cross-sectional study was conducted among 450 continuation phase patients registered at the CCC. The continuation phase was selected to ensure information on practices during the full intensive phase was retrieved, as the intensive phase is the most critical phase of anti-TB treatment. The study population was limited to adults (age ≥ 18 years). Patients determined to be mentally handicapped by The Montreal Cognitive Assessment Scale (MoCA) were excluded from the study sample.^14^ As there was no local data on post-COVID DOT prevalence, the minimum necessary sample size was determined to be 427, where n is the desired sample size, z is the confidence level (95%), p is the anticipated population proportion (50%), d is marginal error (5%), and the non-response rate is 10% for simple random sampling:^15^

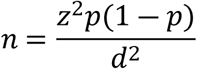

Data was collected via an interviewer-administered questionnaire, and patient files were reviewed for registration information and clinical details. The structured, pre-tested questionnaire was developed in English and translated into Sinhala and Tamil languages. As patient turnout at the CCC is 120-150 per month on average (personal communication), data was collected over four months. All eligible patients present during June 23^rd^ – September 13^th^ 2023 were included. Of the 450 patients interviewed, 444 were attending the CCC and 6 were temporarily residing in the Welikada Prison.

Information gathered included sociodemographic and economic factors, clinical details, knowledge and attitudes about TB, and DOT practices. Standard of living was assessed as suggested by The Demographic and Health Surveys by accessibility of ten amenities (drinking water, toilet facilities, floor and roof of house, electricity, television, radio, telephone, refrigerator, and vehicle) ranked on a 0-2 Likert scale and summed, with ‘High’ (>75th percentile), ‘Medium’ (25th – 75th percentile), or ‘Low’ (<25th percentile).^16^ Physical disability was measured using the Katz Index of Independence in Activities of Daily Living in which performance in each of six functions (bathing, dressing, toileting, transferring, continence, and feeding) is ‘Independent’ (one point) or ‘Dependent’ (zero points), and respondents are classified as ‘Independent’ (≥50th percentile) or ‘Dependent’ (<50th percentile) overall by total score.^17^ Knowledge was assessed by six ‘True’ or ‘False’ statements (on the infectious nature of TB, possibility of death, curability, duration of treatment, consequences of default, and requirement of supervision) with one mark given per correct answer, with ‘Adequate’ (≥4) and ‘Inadequate’ (<4) knowledge judged by the sum of marks. Social support was assessed using the Duke-UNC Functional Social Support Questionnaire in which eight statements on various aspects of support are ranked from 1 (‘Much less than I would like’) to 5 (‘As much as I would like’) and averaged marks determined ‘Inadequate’ (≤3) or ‘Adequate’ (>3).^18^

### Statistical analysis and data management

DOT compliance was defined as the presence of a DOT provider, with patients considered noncompliant if they lacked a DOT provider outside of the facility. Note that this study specifically investigates compliance with DOT–a practice hallmarked by the act of witness–and not TB treatment compliance in general, which extends beyond this definition.

Both descriptive and inferential statistics were used. Univariate, bivariate, and multivariate analyses were performed, and logistic regression was used to identify factors independently associated with presence of a DOT provider after accounting for confounders. P-values < 0.05 were considered to indicate statistical significance. A false discovery rate correction was performed to account for multiple testing, reported as q-values. Distance from each patient’s address to the CCC was determined by Google Maps with the prevailing road structure. All analyses were conducted in R, with figures produced using the package ggplot2.^19,20^

### Ethical considerations

The study has been approved by the Ethics Review Committee of the National Institute of Health Sciences, Ministry of Health, Sri Lanka, which complies with the appropriate guidelines of the Forum of Ethics Review Committees in Sri Lanka (Reference No: NIHS/ERC/23/17R).

All eligible participants were given information about the study and provided the opportunity to ask questions. All patients included were asked to voluntarily participate and have given their informed written consent before impartial witnesses. All data obtained and used for research purposes are kept completely confidential.

## Results

Of the 450 patients interviewed, 117 (26.0%) were DOT compliant and 333 (74.0%) were DOT noncompliant, with compliance defined as having a DOT provider (S1).

### Sociodemographic and economic factors

Most patients were in the most economically productive ages of 30-60 years (61.3%), male (61.1%), Sinhalese (53.1%), and residing within 5 kilometres of the CCC (43.6%). The majority were also married (72.4%), living in extended families (87.3%), had a ‘High’ standard of living score (90.4%), literate (77.8%), and educated up to Advanced Level (42.4%). Patients were often unemployed (67.6%) and earning household income within 60,000-100,000 LKR (37.6%)–well below the 2022 GDP per capita of roughly 1 million LKR.^21^ Most patients experienced lost household income due to their TB diagnosis (94.2%).

Of these factors, patients who reported lost household income due to their diagnosis were significantly associated with poor DOTS compliance (p=0.004), and both male sex (p=0.061) and living alone (p=0.060) showed marginal significance (Table 1).

**Table 1.**
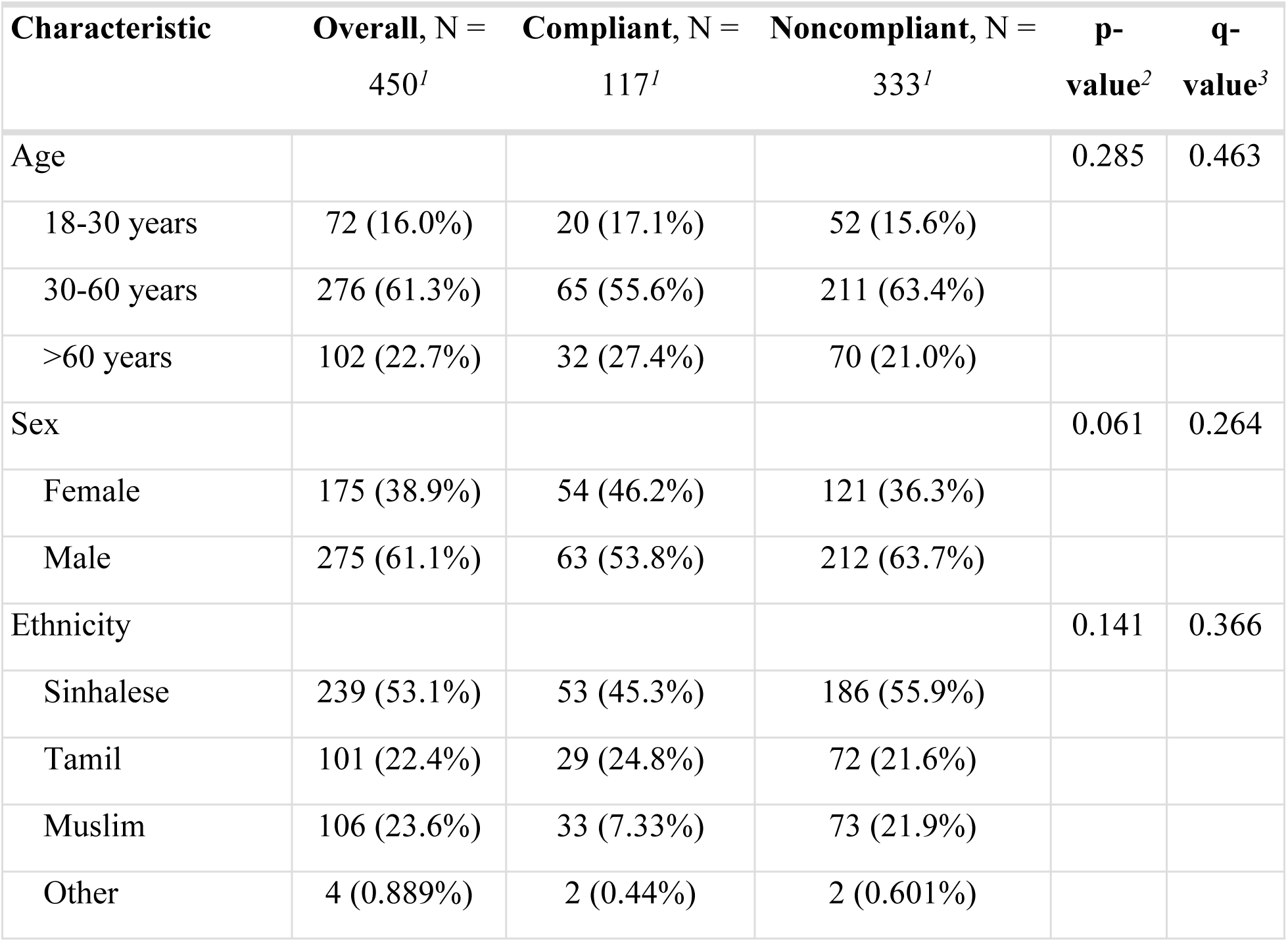
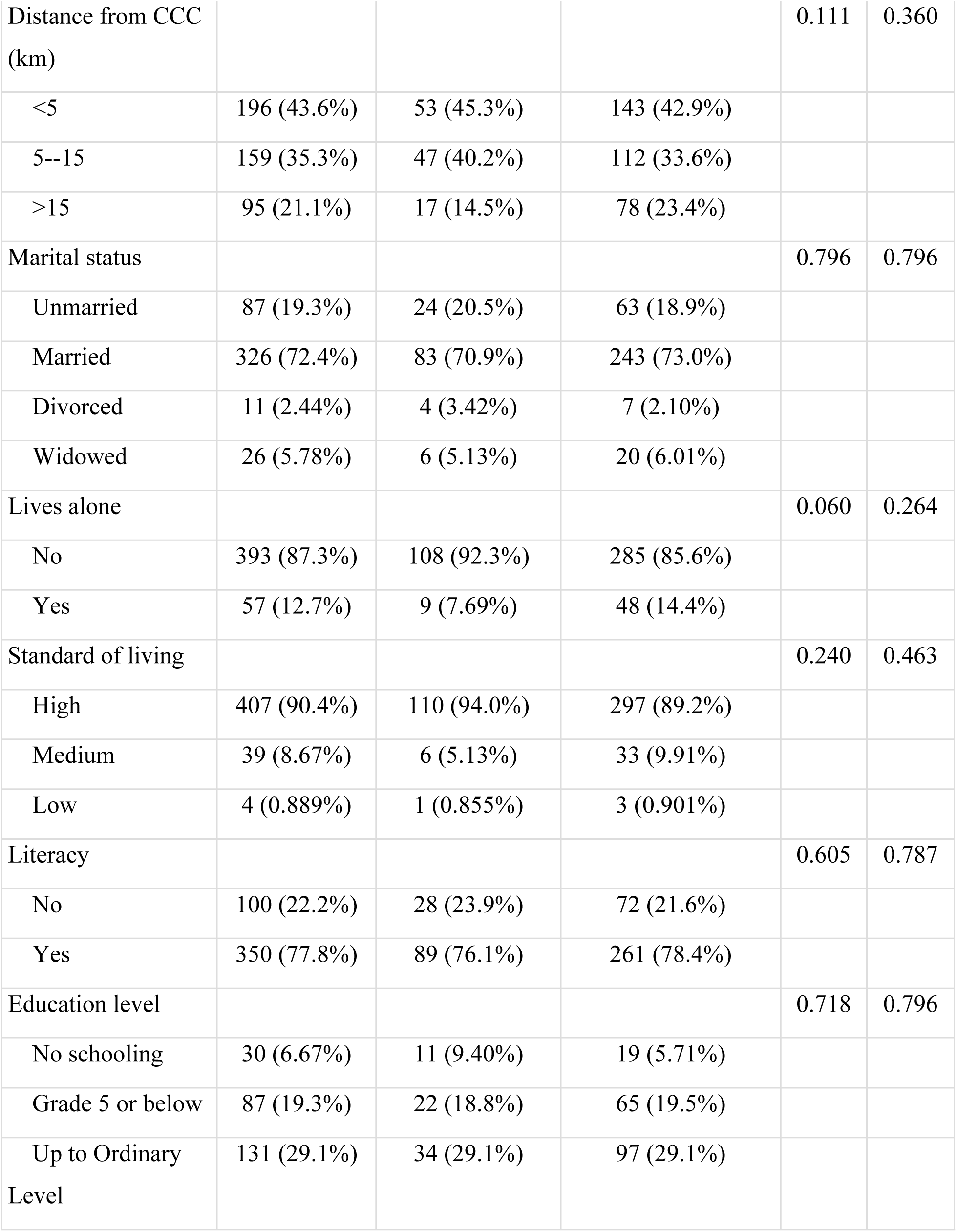
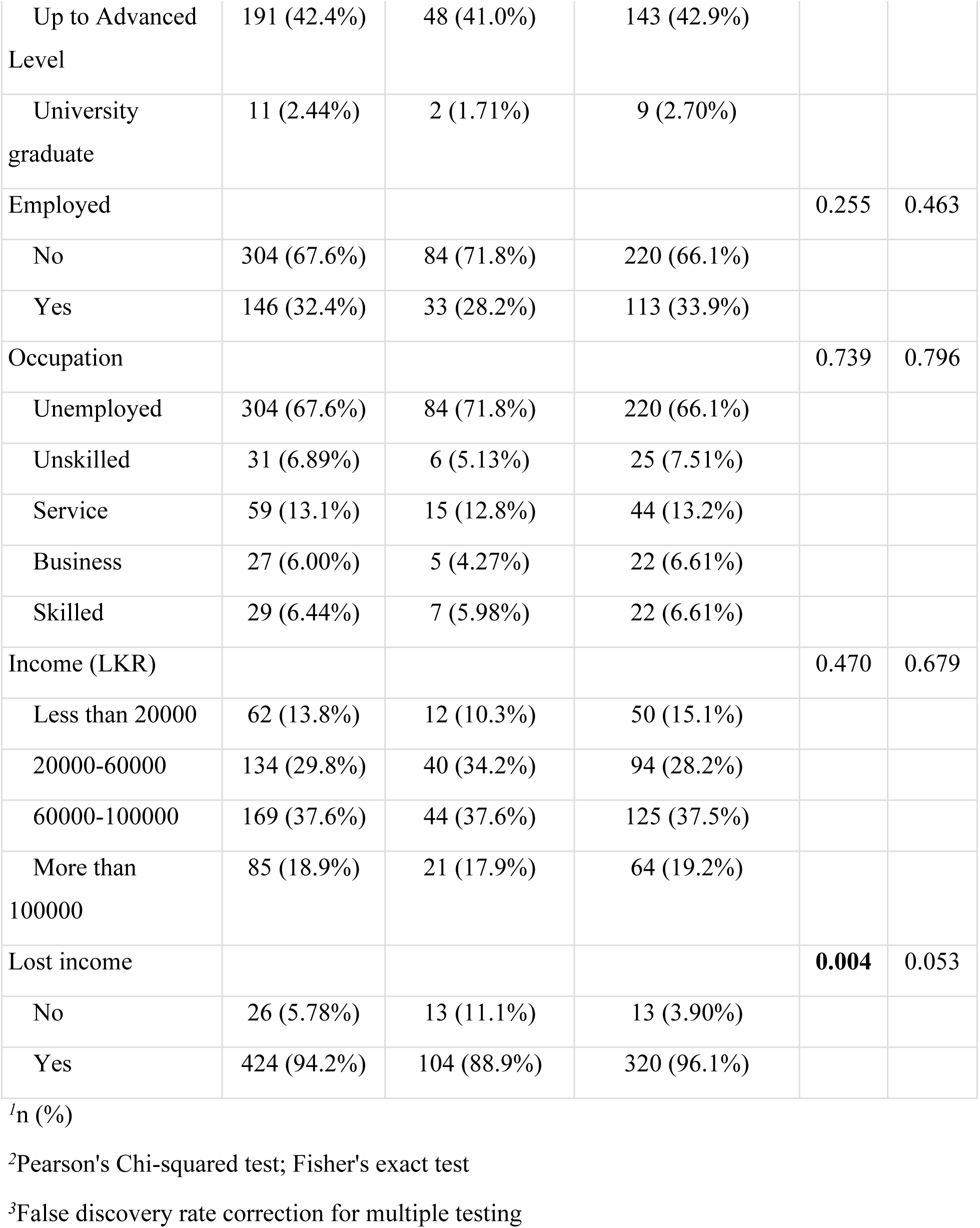
DOT compliance according to sociodemographic and economic factors.

| Characteristic | Overall, N = 450<sup>1</sup> | Compliant, N = 117<sup>1</sup> | Noncompliant, N = 333<sup>1</sup> | p-value<sup>2</sup> | q-value<sup>3</sup> |
| --- | --- | --- | --- | --- | --- |
| Age |  |  |  | 0.285 | 0.463 |
| 18-30 years | 72 (16.0%) | 20 (17.1%) | 52 (15.6%) |  |  |
| 30-60 years | 276 (61.3%) | 65 (55.6%) | 211 (63.4%) |  |  |
| >60 years | 102 (22.7%) | 32 (27.4%) | 70 (21.0%) |  |  |
| Sex |  |  |  | 0.061 | 0.264 |
| Female | 175 (38.9%) | 54 (46.2%) | 121 (36.3%) |  |  |
| Male | 275 (61.1%) | 63 (53.8%) | 212 (63.7%) |  |  |
| Ethnicity |  |  |  | 0.141 | 0.366 |
| Sinhalese | 239 (53.1%) | 53 (45.3%) | 186 (55.9%) |  |  |
| Tamil | 101 (22.4%) | 29 (24.8%) | 72 (21.6%) |  |  |
| Muslim | 106 (23.6%) | 33 (7.33%) | 73 (21.9%) |  |  |
| Other | 4 (0.889%) | 2 (0.44%) | 2 (0.601%) |  |  |
| Distance from CCC (km) |  |  |  | 0.111 | 0.360 |
| <5 | 196 (43.6%) | 53 (45.3%) | 143 (42.9%) |  |  |
| 5--15 | 159 (35.3%) | 47 (40.2%) | 112 (33.6%) |  |  |
| >15 | 95 (21.1%) | 17 (14.5%) | 78 (23.4%) |  |  |
| Marital status |  |  |  | 0.796 | 0.796 |
| Unmarried | 87 (19.3%) | 24 (20.5%) | 63 (18.9%) |  |  |
| Married | 326 (72.4%) | 83 (70.9%) | 243 (73.0%) |  |  |
| Divorced | 11 (2.44%) | 4 (3.42%) | 7 (2.10%) |  |  |
| Widowed | 26 (5.78%) | 6 (5.13%) | 20 (6.01%) |  |  |
| Lives alone |  |  |  | 0.060 | 0.264 |
| No | 393 (87.3%) | 108 (92.3%) | 285 (85.6%) |  |  |
| Yes | 57 (12.7%) | 9 (7.69%) | 48 (14.4%) |  |  |
| Standard of living |  |  |  | 0.240 | 0.463 |
| High | 407 (90.4%) | 110 (94.0%) | 297 (89.2%) |  |  |
| Medium | 39 (8.67%) | 6 (5.13%) | 33 (9.91%) |  |  |
| Low | 4 (0.889%) | 1 (0.855%) | 3 (0.901%) |  |  |
| Literacy |  |  |  | 0.605 | 0.787 |
| No | 100 (22.2%) | 28 (23.9%) | 72 (21.6%) |  |  |
| Yes | 350 (77.8%) | 89 (76.1%) | 261 (78.4%) |  |  |
| Education level |  |  |  | 0.718 | 0.796 |
| No schooling | 30 (6.67%) | 11 (9.40%) | 19 (5.71%) |  |  |
| Grade 5 or below | 87 (19.3%) | 22 (18.8%) | 65 (19.5%) |  |  |
| Up to Ordinary Level | 131 (29.1%) | 34 (29.1%) | 97 (29.1%) |  |  |
| Up to Advanced Level | 191 (42.4%) | 48 (41.0%) | 143 (42.9%) |  |  |
| University graduate | 11 (2.44%) | 2 (1.71%) | 9 (2.70%) |  |  |
| Employed |  |  |  | 0.255 | 0.463 |
| No | 304 (67.6%) | 84 (71.8%) | 220 (66.1%) |  |  |
| Yes | 146 (32.4%) | 33 (28.2%) | 113 (33.9%) |  |  |
| Occupation |  |  |  | 0.739 | 0.796 |
| Unemployed | 304 (67.6%) | 84 (71.8%) | 220 (66.1%) |  |  |
| Unskilled | 31 (6.89%) | 6 (5.13%) | 25 (7.51%) |  |  |
| Service | 59 (13.1%) | 15 (12.8%) | 44 (13.2%) |  |  |
| Business | 27 (6.00%) | 5 (4.27%) | 22 (6.61%) |  |  |
| Skilled | 29 (6.44%) | 7 (5.98%) | 22 (6.61%) |  |  |
| Income (LKR) |  |  |  | 0.470 | 0.679 |
| Less than 20000 | 62 (13.8%) | 12 (10.3%) | 50 (15.1%) |  |  |
| 20000-60000 | 134 (29.8%) | 40 (34.2%) | 94 (28.2%) |  |  |
| 60000-100000 | 169 (37.6%) | 44 (37.6%) | 125 (37.5%) |  |  |
| More than 100000 | 85 (18.9%) | 21 (17.9%) | 64 (19.2%) |  |  |
| Lost income |  |  |  | **0.004** | 0.053 |
| No | 26 (5.78%) | 13 (11.1%) | 13 (3.90%) |  |  |
| Yes | 424 (94.2%) | 104 (88.9%) | 320 (96.1%) |  |  |

<sup>1</sup>n (%)
<sup>2</sup>Pearson's Chi-squared test; Fisher's exact test
<sup>3</sup>False discovery rate correction for multiple testing

### Clinical information

The majority of patients were new (94.2%) and diagnosed with PTB (76.0%). Most had been bacteriologically confirmed (67.8%)–of these, most had successful sputum conversion at follow-up smear testing (64.9%). A slight majority had comorbidities (52.7%), of which diabetes mellitus was the most common (39.6%) (S3). Most were non-smokers (85.1%), non-alcoholics (93.6%), and did not use illicit drugs (97.1%). The majority were determined to be ‘Independent’ in function (96.7%), were of healthy weight upon initial diagnosis (49.8%), and experienced an increase in BMI over the duration of treatment thus far (75.1%).

Both illicit drug use (p=0.007) and physical disability (p=0.005) showed significant association with DOTS compliance (Table 2). Patients with kidney disease displayed marginal significance (p=0.075), and those who reported experiencing another chronic illness not mentioned showed significance (p=0.027), although it should be acknowledged that the latter encompasses a wide range of conditions (S3). Interestingly, presence of a DOT provider appeared to have no bearing on other clinical parameters, including sputum results and change in BMI.

**Table 2.**
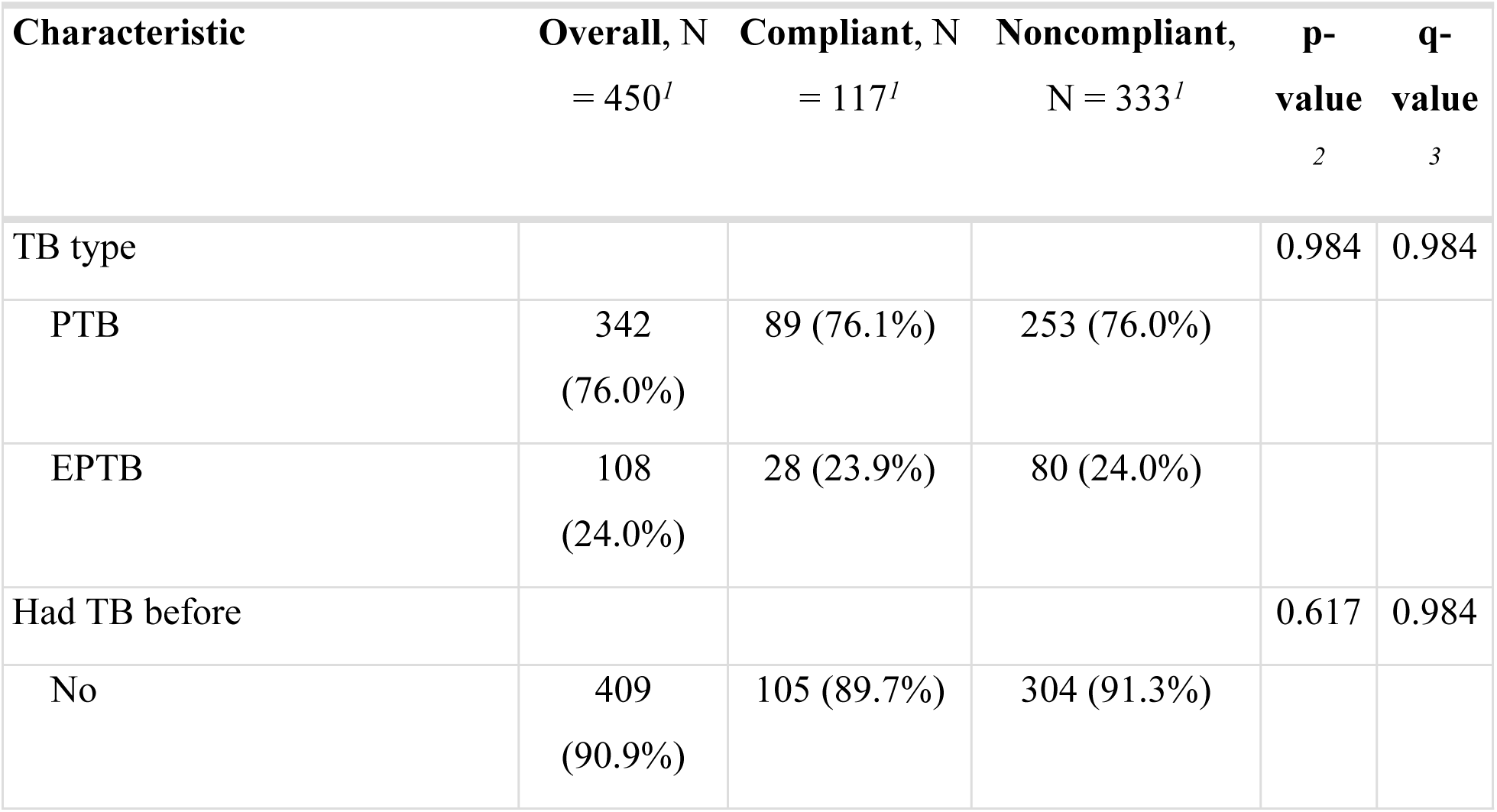
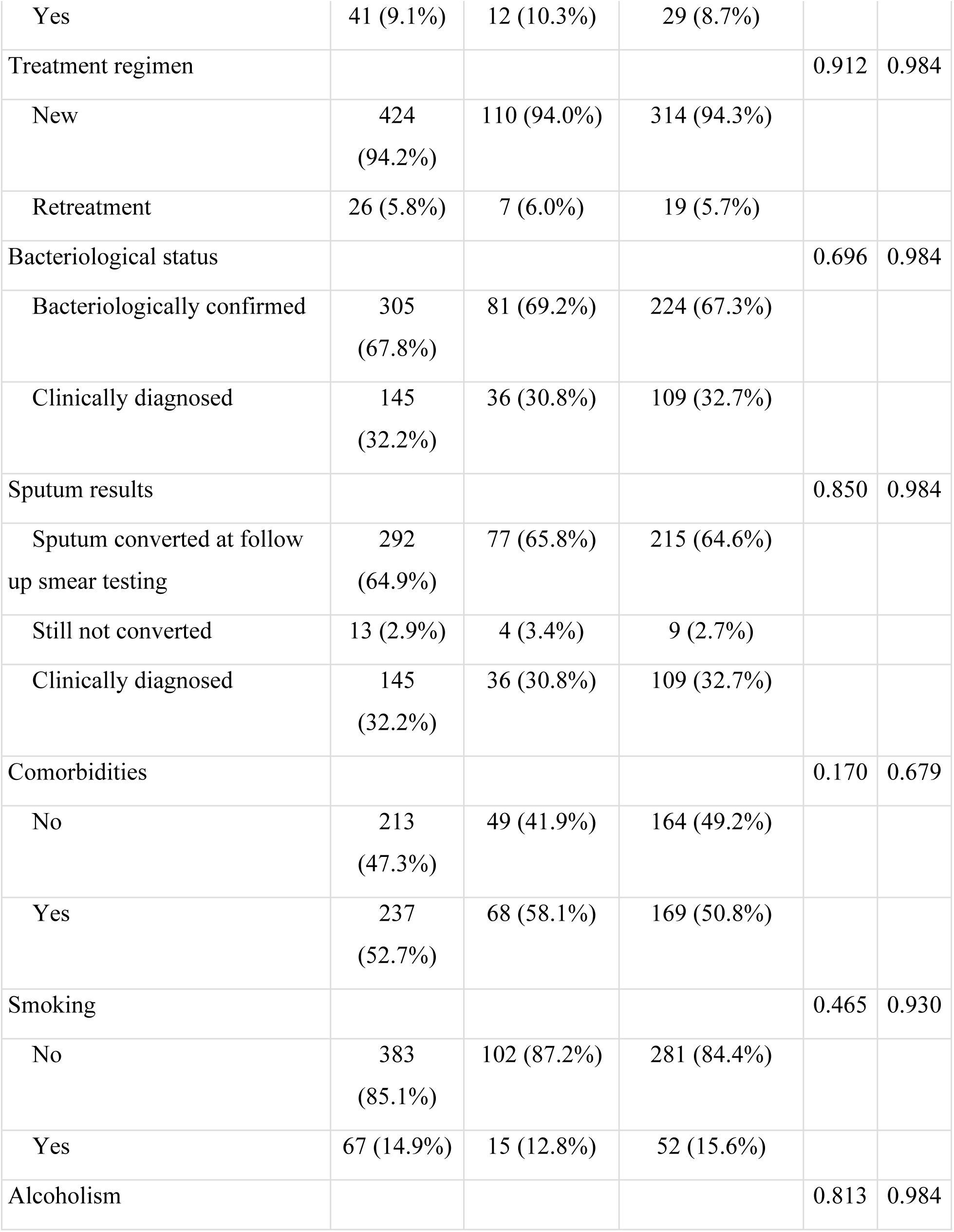
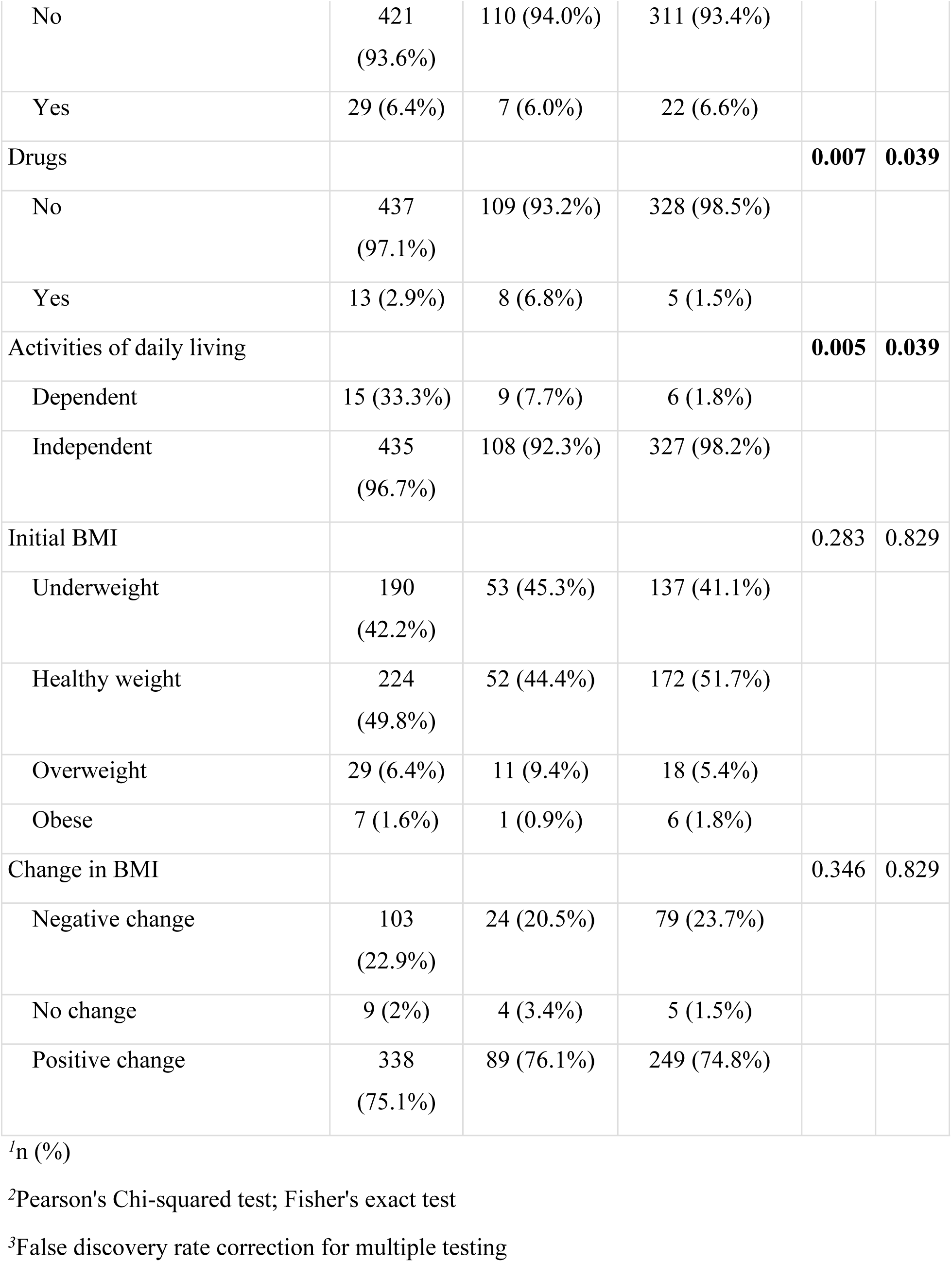
DOT compliance according to clinical information.

| Characteristic | Overall, N<br>= 450<sup>1</sup> | Compliant, N<br>= 117<sup>1</sup> | Noncompliant,<br>N = 333<sup>1</sup> | p-value<br>2 | q-value<br>3 |
| --- | --- | --- | --- | --- | --- |
| TB type |  |  |  | 0.984 | 0.984 |
| PTB | 342<br>(76.0%) | 89 (76.1%) | 253 (76.0%) |  |  |
| EPTB | 108<br>(24.0%) | 28 (23.9%) | 80 (24.0%) |  |  |
| Had TB before |  |  |  | 0.617 | 0.984 |
| No | 409<br>(90.9%) | 105 (89.7%) | 304 (91.3%) |  |  |
| Yes | 41 (9.1%) | 12 (10.3%) | 29 (8.7%) |  |  |
| Treatment regimen |  |  |  | 0.912 | 0.984 |
| New | 424<br>(94.2%) | 110 (94.0%) | 314 (94.3%) |  |  |
| Retreatment | 26 (5.8%) | 7 (6.0%) | 19 (5.7%) |  |  |
| Bacteriological status |  |  |  | 0.696 | 0.984 |
| Bacteriologically confirmed | 305<br>(67.8%) | 81 (69.2%) | 224 (67.3%) |  |  |
| Clinically diagnosed | 145<br>(32.2%) | 36 (30.8%) | 109 (32.7%) |  |  |
| Sputum results |  |  |  | 0.850 | 0.984 |
| Sputum converted at follow<br>up smear testing | 292<br>(64.9%) | 77 (65.8%) | 215 (64.6%) |  |  |
| Still not converted | 13 (2.9%) | 4 (3.4%) | 9 (2.7%) |  |  |
| Clinically diagnosed | 145<br>(32.2%) | 36 (30.8%) | 109 (32.7%) |  |  |
| Comorbidities |  |  |  | 0.170 | 0.679 |
| No | 213<br>(47.3%) | 49 (41.9%) | 164 (49.2%) |  |  |
| Yes | 237<br>(52.7%) | 68 (58.1%) | 169 (50.8%) |  |  |
| Smoking |  |  |  | 0.465 | 0.930 |
| No | 383<br>(85.1%) | 102 (87.2%) | 281 (84.4%) |  |  |
| Yes | 67 (14.9%) | 15 (12.8%) | 52 (15.6%) |  |  |
| Alcoholism |  |  |  | 0.813 | 0.984 |
| No | 421<br>(93.6%) | 110 (94.0%) | 311 (93.4%) |  |  |
| Yes | 29 (6.4%) | 7 (6.0%) | 22 (6.6%) |  |  |
| Drugs |  |  |  | **0.007** | **0.039** |
| No | 437<br>(97.1%) | 109 (93.2%) | 328 (98.5%) |  |  |
| Yes | 13 (2.9%) | 8 (6.8%) | 5 (1.5%) |  |  |
| Activities of daily living |  |  |  | **0.005** | **0.039** |
| Dependent | 15 (33.3%) | 9 (7.7%) | 6 (1.8%) |  |  |
| Independent | 435<br>(96.7%) | 108 (92.3%) | 327 (98.2%) |  |  |
| Initial BMI |  |  |  | 0.283 | 0.829 |
| Underweight | 190<br>(42.2%) | 53 (45.3%) | 137 (41.1%) |  |  |
| Healthy weight | 224<br>(49.8%) | 52 (44.4%) | 172 (51.7%) |  |  |
| Overweight | 29 (6.4%) | 11 (9.4%) | 18 (5.4%) |  |  |
| Obese | 7 (1.6%) | 1 (0.9%) | 6 (1.8%) |  |  |
| Change in BMI |  |  |  | 0.346 | 0.829 |
| Negative change | 103<br>(22.9%) | 24 (20.5%) | 79 (23.7%) |  |  |
| No change | 9 (2%) | 4 (3.4%) | 5 (1.5%) |  |  |
| Positive change | 338<br>(75.1%) | 89 (76.1%) | 249 (74.8%) |  |  |

<sup>1</sup>n (%)
<sup>2</sup>Pearson's Chi-squared test; Fisher's exact test
<sup>3</sup>False discovery rate correction for multiple testing

### Knowledge and attitudes

Patients demonstrated adequate overall knowledge (94.2%), as well as adequate specific knowledge of the disease’s infectious nature (79.3%), possibility of death (87.8%), curability (97.6%), treatment duration (88.4%), consequences of default (92.9%), and treatment supervision (83.7%). Most reported not experiencing stigma (84.7%) and receiving adequate social support (82.4%).

While overall knowledge score showed no association with DOT compliance, both specific knowledge of the curability of the disease (p=0.039) and the necessity of a DOT provider itself (p=0.009) significantly affected DOT compliance (Table 3). When analysed in relation to sociodemographic and economic factors, knowledge of the supervision requirement showed a strong association with living alone (p=0.027), standard of living score (p=0.003), and income (p<0.001) (S4). Regarding attitudes, stigma had no bearing on the likelihood of having a DOT provider, while social support score displayed a strong correlation (p=0.003).

**Table 3.**
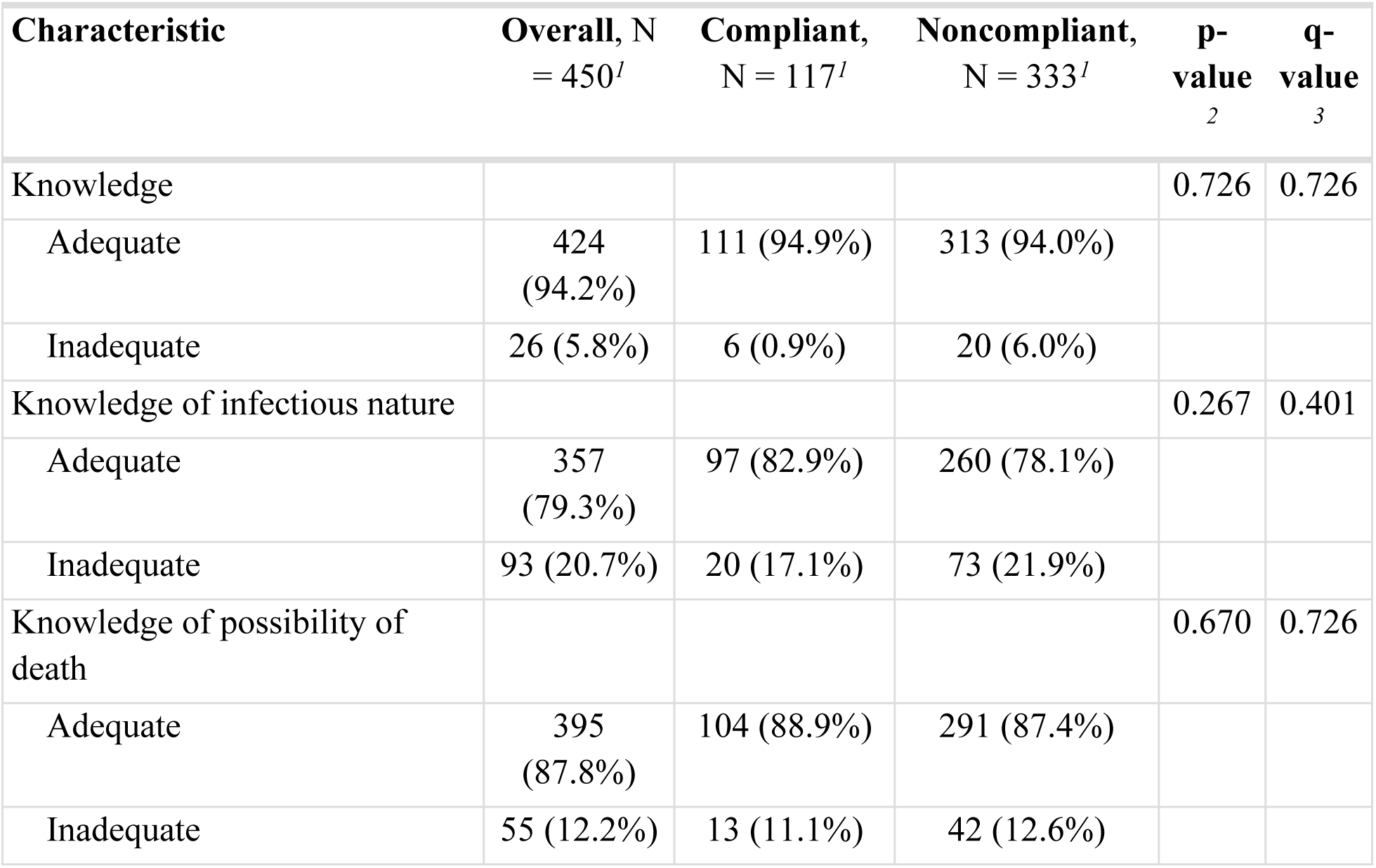
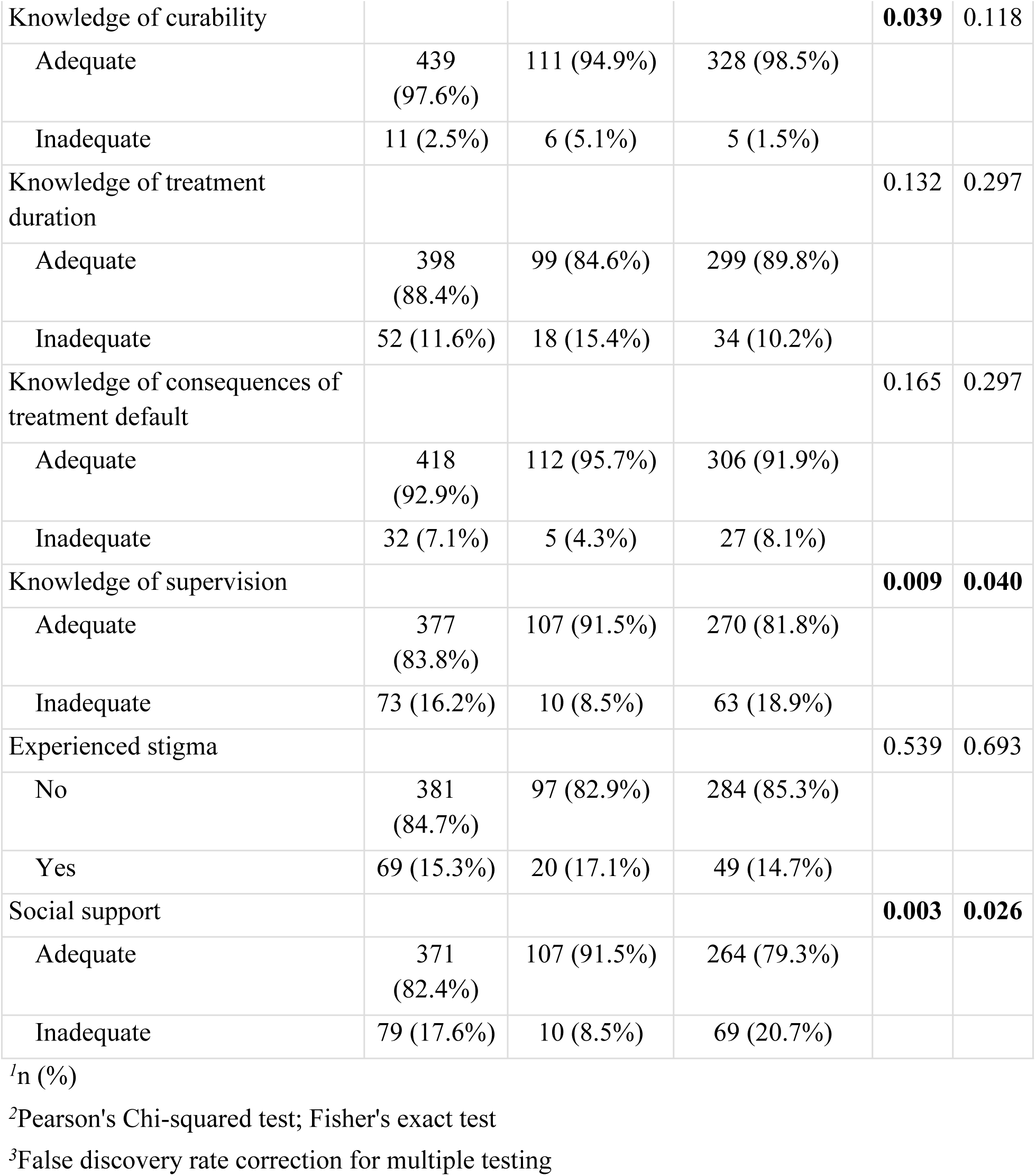
DOT compliance according to knowledge and attitudes.

| Characteristic | Overall, N<br>= 450<sup>1</sup> | Compliant,<br>N = 117<sup>1</sup> | Noncompliant,<br>N = 333<sup>1</sup> | p-value<br>2 | q-value<br>3 |
| --- | --- | --- | --- | --- | --- |
| Knowledge |  |  |  | 0.726 | 0.726 |
| Adequate | 424<br>(94.2%) | 111 (94.9%) | 313 (94.0%) |  |  |
| Inadequate | 26 (5.8%) | 6 (0.9%) | 20 (6.0%) |  |  |
| Knowledge of infectious nature |  |  |  | 0.267 | 0.401 |
| Adequate | 357<br>(79.3%) | 97 (82.9%) | 260 (78.1%) |  |  |
| Inadequate | 93 (20.7%) | 20 (17.1%) | 73 (21.9%) |  |  |
| Knowledge of possibility of death |  |  |  | 0.670 | 0.726 |
| Adequate | 395<br>(87.8%) | 104 (88.9%) | 291 (87.4%) |  |  |
| Inadequate | 55 (12.2%) | 13 (11.1%) | 42 (12.6%) |  |  |
| Knowledge of curability |  |  |  | **0.039** | 0.118 |
| Adequate | 439<br>(97.6%) | 111 (94.9%) | 328 (98.5%) |  |  |
| Inadequate | 11 (2.5%) | 6 (5.1%) | 5 (1.5%) |  |  |
| Knowledge of treatment duration |  |  |  | 0.132 | 0.297 |
| Adequate | 398<br>(88.4%) | 99 (84.6%) | 299 (89.8%) |  |  |
| Inadequate | 52 (11.6%) | 18 (15.4%) | 34 (10.2%) |  |  |
| Knowledge of consequences of treatment default |  |  |  | 0.165 | 0.297 |
| Adequate | 418<br>(92.9%) | 112 (95.7%) | 306 (91.9%) |  |  |
| Inadequate | 32 (7.1%) | 5 (4.3%) | 27 (8.1%) |  |  |
| Knowledge of supervision |  |  |  | **0.009** | **0.040** |
| Adequate | 377<br>(83.8%) | 107 (91.5%) | 270 (81.8%) |  |  |
| Inadequate | 73 (16.2%) | 10 (8.5%) | 63 (18.9%) |  |  |
| Experienced stigma |  |  |  | 0.539 | 0.693 |
| No | 381<br>(84.7%) | 97 (82.9%) | 284 (85.3%) |  |  |
| Yes | 69 (15.3%) | 20 (17.1%) | 49 (14.7%) |  |  |
| Social support |  |  |  | **0.003** | **0.026** |
| Adequate | 371<br>(82.4%) | 107 (91.5%) | 264 (79.3%) |  |  |
| Inadequate | 79 (17.6%) | 10 (8.5%) | 69 (20.7%) |  |  |

<sup>1</sup>n (%)
<sup>2</sup>Pearson's Chi-squared test; Fisher's exact test
<sup>3</sup>False discovery rate correction for multiple testing

### DOTS details and feedback

Most patients reported no missed doses (87.1%)–of those who did, most missed 1-week’s worth or less (11.6%). A majority experienced no adverse reaction to the anti-TB drugs (55.3%).

Interestingly, neither missed doses nor amount of doses missed showed any relation to DOTS compliance (Table 4). Among the patients with a DOT provider, the majority reported a daily treatment frequency (76.1%) under the supervision of a family member (87.2%) with whom they were satisfied (98.3%) (S5). Most patients without a DOT provider cited doing so due to personal preference for self-administered therapy (60.7%); however, it is notable that 28 patients were simply unaware of the DOT supervision requirement itself (S6). Of the patients who reported missing doses, most (53.4%) sought advice to resume regular treatment, usually from the chest clinic (80.6%) (S7). A larger majority of those who reported experiencing an adverse reaction to treatment sought advice (88.1%), often from the chest clinic as well (92.7%) (S8). While the majority of both compliant and noncompliant patients preferred to meet their DOT provider in person (64.7%), many reported being open to a photograph (57.1%) or video call (50.0%) as an alternative (S9).

**Table 4.** DOT compliance according to treatment details.

| **Characteristic** | **Overall, N**<br>**= 450<sup>1</sup>** | **Compliant, N**<br>**= 117<sup>1</sup>** | **Noncompliant,**<br>**N = 333<sup>1</sup>** | **p-**<br>**value**<br>**2** | **q-**<br>**value**<br>**3** |
| --- | --- | --- | --- | --- | --- |
| Missed doses |  |  |  | 0.349 | 0.419 |
| No | 392<br>(87.1%) | 99 (84.6%) | 293 (88.0%) |  |  |
| Yes | 58 (12.9%) | 18 (15.4%) | 40 (12.0%) |  |  |
| Amount of doses<br>missed |  |  |  | 0.147 | 0.419 |
| 1 week or less | 52 (11.6%) | 15 (12.8%) | 37 (11.1%) |  |  |
| 1-3 weeks | 2 (0.4%) | 2 (1.7%) | 0 (0.0%) |  |  |
| >3 weeks | 4 (0.9%) | 1 (0.9%) | 3 (0.9%) |  |  |
| None | 392<br>(87.1%) | 99 (84.6%) | 293 (88.0%) |  |  |
| Adverse reaction |  |  |  | 0.419 | 0.419 |
| No | 249<br>(55.3%) | 61 (52.1%) | 188 (56.5%) |  |  |
| Yes | 201<br>(44.7%) | 56 (47.9%) | 145 (43.5%) |  |  |

<sup>1</sup>n (%)<sup>2</sup>Pearson's Chi-squared test; Fisher's exact test<sup>3</sup>False discovery rate correction for multiple testing

### Factors associated with DOTS compliance

Sex, lost household income, illicit drug use, physical disability, social support, and knowledge of curability and supervision showed significant association with DOTS compliance when tested in the presence of other predictors (Table 5). Patients who were male (OR 0.57; 95% CI 0.36-0.91), reported lost income (OR 0.36; 95% CI 0.15-0.89), had no physical disability (OR 0.17; 95% CI 0.05-0.54), experienced inadequate social support (OR 0.27; 95% CI 0.11-0.61), or were aware of the curability of TB (OR 0.26; 95% CI 0.06-1.00) were more likely to lack a DOT provider. Meanwhile, patients who were aware of the requirement of supervision (OR 3.77; 95% CI 1.73-9.27) or who frequently used illicit drugs (OR 14.4; 95% CI 3.75-62.3) were more likely to have a DOT provider. However, this relationship between DOTS compliance and illicit drugs is confounded by 6 of the 13 frequent drug users residing in the Welikada Prison, where DOT supervision is enforced. Beyond this study, this model can also be used to predict noncompliance in patients, the overall misclassification rate of which is 23.8%.

**Table 5.**
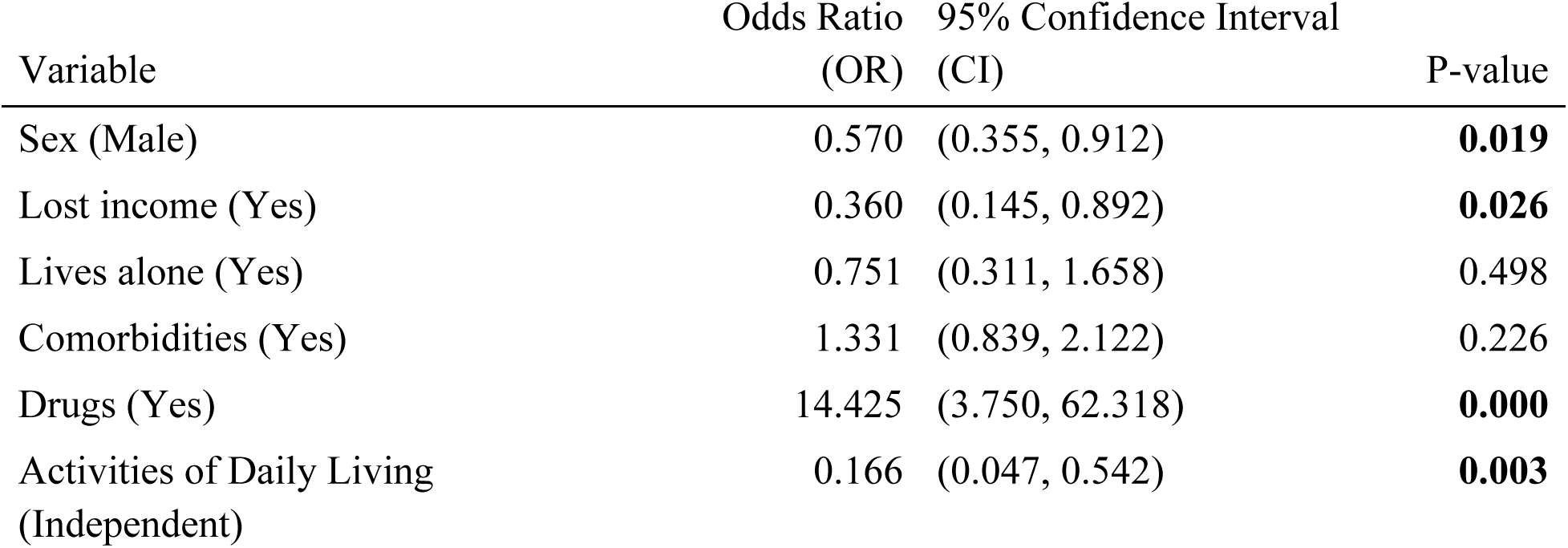
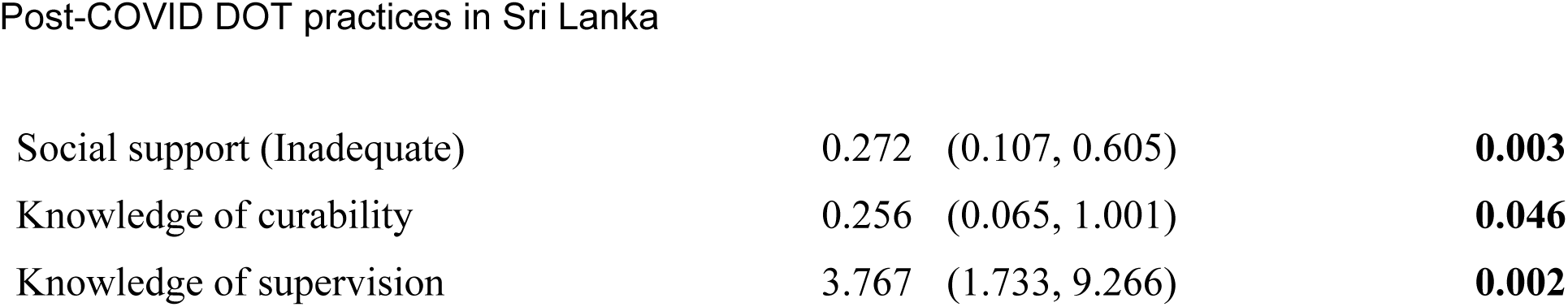
Multiple logistic regression of predictors of DOT compliance.

## Discussion

This is the first cross-sectional descriptive study on DOT post-COVID-19 in Sri Lanka. It contributes to discussion of alternatives in resource-limited circumstances in which facility-based DOT is not feasible.

In this study, DOT compliance refers specifically to presence of a DOT provider, and should not be conflated with compliance to TB treatment, as most patients who were noncompliant with the former were still compliant with the latter. Defined as the lack of a DOT provider, we report a DOT noncompliance rate of 74.0% within our study population. This drop to only about one fourth (26.0%) adhering to DOT protocol is drastic compared to the 100% DOT by healthcare providers pre-COVID. For patients still adherent to DOT protocol, the main provider identified was a family member (87.2%), followed by a healthcare worker (9.4%) or neighbor (1.7%). However, the observed DOT coverage still appears to be higher than that reported in South Africa and India, possibly due to comparatively stronger healthcare infrastructure within the Colombo district.^22,23^

Most patients had resorted to unauthorised self-administered therapy (SAT), but we observed that DOT supervision or lack thereof had no statistically significant effect on clinical outcomes such as sputum conversion, change in BMI, or missed doses. Although the Kalutara District also reported a high DOT noncompliance rate of 61.9% in 2020, other studies show significant association between such noncompliance and missed doses, which contradicts our findings.^24,25^ Attributes of our noncompliant patients carry over from those which characterise Colombo’s TB patient population itself: largely unemployed and of low socioeconomic status, with few having achieved higher education and engaging in skilled labor.^26^ While the DOT provider of most compliant patients was a family member with whom they were satisfied, most patients without a DOT provider preferred SAT. The CCC remains patients’ primary source of advice regarding missed doses and adverse reactions.

Factors independently associated with noncompliance were male sex, lost income due to diagnosis, no physical disability, inadequate social support, and awareness of curability. Patients informed of the DOT strategy were more likely to be compliant, underscoring the importance of proper health education as several noncompliant patients were simply unaware of the supervision requirement itself. Further, adequate social support was associated with the presence of a DOT provider, indicating an importance of community-centered care in anti-TB treatment.

Interestingly, the proportion of patients with no missed doses who were DOT noncompliant (defined as lacking a DOT provider) was comparable to that of patients who were DOT compliant (defined as having a DOT provider), suggesting a greater significance of patient self-reliance within the study population. Under the family DOT model assessed, factors formerly associated with utilisation of in-person DOT services such as age, occupation, income, and residence were not significantly associated with DOT compliance, likely because travel to the CCC is irrelevant to DOT services provided at home.^27^

Limitations of the study include patients’ self-reporting and the pitfalls of direct questioning, as well as the selection bias of interviewing patients already seeking medical attention. We additionally note that as our standard of living survey tool equates basic amenities such as drinking water, toilet facilities, and a roof to accessories such as a vehicle or radio, patients’ standards of living may be artificially inflated. Concerns regarding the underlying conditions for Chi-squared test procedures are also raised due to small sample sizes in some categories.

More robust anti-TB treatment can also be achieved by engaging partners beyond the health sector.^1^ Amidst escalating food insecurity and household expenses, government interventions must better support lost income to break the cyclical relationship between low socioeconomic status and TB.^12^ Education about DOT supervision must be improved. Further, DOT compliance has been shown to be associated with motivation from friends, family, and healthcare workers.^28^ Policymakers must aggressively combat stigma of diagnosis, as family support is linked to treatment success and inadequate social support was identified as a cause of noncompliance.^29^ For instance, appointing family DOT providers who may be unsupportive risks impeding DOT practices. With the CCC as patients’ primary source of guidance, reported poor interactions with staff must be amended.^30^ While most patients still prefer to meet their DOT provider, technological assistance through photographs and video calls are future alternatives to explore.

This study suggests SAT as a viable DOT alternative under resource-limited circumstances. This finding parallels other studies which similarly report no statistically significant difference between patients’ DOT and SAT clinical outcomes.^29,31^ SAT has already been implemented in remote, resource-limited, or conflict-affected settings around the globe in which facility-based DOT is not feasible.^29,32^ Moreover, community-based DOT has been shown to improve TB treatment outcomes.^33^ High SAT success rates have been observed provided that risk factors are considered when determining treatment delivery models; however, with a greater onus on the individual, ongoing health education and patient empowerment becomes all the more essential.^29^

## Conclusion

Overall, our study reveals widespread noncompliance with the DOT practices that are advised by the NPTCCD, largely due to a common preference for SAT. However, SAT may be a possible alternative to DOT as clinical outcomes show no significant difference between the two approaches, provided sufficient patient and community health education. DOT strategies must be thoughtfully revised to align with post-COVID realities, as the impact of the country’s social upheaval on the practice has been transformative.

## Data Availability

Data cannot be shared publicly because they contain private patient information, including clinical details and addresses. A de-identified dataset with patient addresses removed is freely available from the authors upon reasonable request. Access to the full dataset, including restricted information, is available from the Ethics Review Committee, National Institute of Health Sciences (P.O.Box 28, Matugama Rd, Nagoda, Kalutara, Sri Lanka Telephone: +94-34-2222264 ext 219) for researchers who meet the criteria for access to confidential data.

## Acknowledgements

The authors thank Mahathi Athreya of Amherst College’s Department of Mathematics & Statistics for assistance with data analysis, as well as the NPTCCD officials and CCC staff for their logistical support and guidance–namely Dr. F.M. Jamaldeen and Dr. P.V.W. Senadheera, who served as impartial witnesses in the clinic and prison, respectively. The views expressed in this publication are those of the author(s) and not necessarily those of the NPTCCD.

